# Temporal trends of hepatitis C virus transmission in internally displaced people who inject drugs in Odessa, Ukraine

**DOI:** 10.1101/2021.12.28.21268494

**Authors:** Anna Yakovleva, Ganna Kovalenko, Matthew Redlinger, Pavlo Smyrnov, Olga Tymets, Ania Korobchuk, Lyudmyla Kotlyk, Ganna Kolodyazeva, Ganna Podolina, Svetlana Chernyavskaya, Steffanie A. Strathdee, Samuel R Friedman, Ian Goodfellow, Joel O. Wertheim, Eric Bortz, Luke Meredith, Tetyana I Vasylyeva

## Abstract

**Background:** Due to practical challenges associated with genetic sequencing in low resource environments, the burden of hepatitis C virus (HCV) in forcibly displaced people is understudied. We examined the use of field-applicable HCV sequencing methods and phylogenetic analysis to determine HCV transmission dynamics in internally displaced people who inject drugs (IDPWID) due to war in eastern Ukraine.

**Methods:** In this cross-sectional study, we used modified respondent-driven sampling to recruit IDPWID who have settled in Odessa, Ukraine. We generated partial and near full length genome (NFLG) HCV sequences using Oxford Nanopore MinION in a simulated field environment. Maximum likelihood and Bayesian methods were used to establish phylodynamic relationships.

**Findings:** Between June and September 2020, we collected epidemiological data and whole blood samples from 164 IDPWID. Rapid testing identified an HIV, anti-HCV, and HIV/HCV co-infection prevalence of 39%, 67.7%, and 31.1%, respectively. We generated 57 partial or NFLG HCV sequences and identified eight transmission clusters, of which at least two originated within a year and a half post-migration. Unstable housing post-migration was associated with more reports of injection drug use in the past 30 days (p=0.048).

**Interpretation:** Local generation of viral sequencing data and phylogenetic analysis in rapidly changing low-resource environments, such as those faced by forcibly displaced people, can inform timely adaptation of prevention and treatment.

**Funding:** Oxford University John Fell Fund, ISS Wellcome Trust, Branco Weiss Fellowship.

## Background

Globally, 82.4 million people have been forcibly displaced following economic, political, and climate instability, which is almost twice as high as two decades earlier [1]. By 2050, a further 143 million people may become internally displaced because of climate change [2]. Internally displaced persons (IDPs, people who do not cross international borders) constitute the majority of forcibly displaced people: currently, there are 48 million IDPs residing in low- and middle-income countries, 85% of whom have been displaced due to conflict and violence [1].

Forced migration is associated with worse health outcomes in migrant compared to the autochthonous population, attributed to increased vulnerabilities and hardships experienced throughout the migration process, which are reinforced by structural inequalities, discrimination, and gender-based violence [3]. The burden of otherwise preventable and controllable infectious diseases is often high amongst IDPs, due at least in part to the collapse of local health systems and infrastructure [4].

Unstable housing post-migration, which broadly encompasses settlement in refugee camps, homelessness, residing in overcrowded homes, as well as with friends and relatives, has been implicated in increased risk of acquiring infections such as HIV post-migration [5]. This relationship is likely to be similar for hepatitis C virus (HCV) infections and might be exacerbated by risk factors such as injection drug use (IDU) [6].

As of 2021, Ukraine has the largest IDPs population in Europe: over 1.4 million people have been internally displaced and over 14,000 killed since the beginning of the war in the Donbass region (which includes Donetsk and Lugansk administrative regions) in eastern Ukraine and the annexation of Crimea in 2014 [7]; 38,600 of IDPs settled in Odessa in southern Ukraine. Concurrently, Ukraine has a high prevalence of IDU, which has caused the second largest HCV epidemic in Europe, following the Russian Federation [8, 9]. National HCV seroprevalence in PWID is 56%: lower amongst PWID in Odessa (47.5%) and Crimea (25.7% – 50.1%) compared to Donetsk (55.2%) and Lugansk (58.4%) [10]. In 2020, national HCV treatment uptake had risen modestly from 0.11% in 2013 to 6.5% in 2020 – an important step towards HCV elimination since treatment can successfully cure the disease in >80% of cases [11] - but only 10-20% of those treated were PWID [12]. Although no data on HCV epidemiology in Ukrainian internally displaced PWID (IDPWID) is currently available, IDPs are likely to experience additional barriers in accessing appropriate interventions and treatment options compared to non-displaced populations [4].

Molecular epidemiology, the study of how epidemiological and evolutionary factors shape viral phylogenies [13], is increasingly utilised in public health settings due to the widespread availability of pathogen genetic sequences and has been used to identify HCV transmission clusters, thus improving our understanding of infection transmission chains [14, 15]. We previously used molecular epidemiology methods to show that internal displacement following war in eastern Ukraine triggered a geographic redistribution of existing HIV infections: migration from high prevalence regions in the east to lower prevalence regions in the west [16]. Similar analyses were not possible for HCV due to a lack of available genetic sequence data, despite Ukraine having one of the highest HCV burdens in the world [9].

Global efforts to reinforce molecular epidemiology-based surveillance of chronic viral infections, such as HIV and HCV, in the context of forced migration are hindered by the scarcity of available pathogen sequencing data, primarily due to practical laboratory limitations concerning hard to reach mobile populations. Recent advances in portable molecular biology tools applicable in low-resource environments, such as Nanopore® sequencing technology, have enabled molecular epidemiological characterisation of other viral pathogens such as Ebola and rabies in the field [17, 18]. Here we demonstrate Nanopore based partial and near full length genome (NFLG) sequencing of HCV clinical samples, which is directly applicable to sequencing in the field. We analysed locally generated sequencing data using phylogenetics to describe HCV transmission dynamics in IDPWID residing in Odessa, Ukraine. By combining our findings with epidemiological data, we reveal opportunities for improved targeting of local preventative interventions and infection management strategies. Overall, this work expands the potential of molecular epidemiology to the study of the burden of HCV in low-resource and rapidly changing environments, such as those faced by forcibly displaced people.

## Methods

### Study design and participants

This study was approved by the University of Oxford Tropical Research Ethical Committee (Reference: 530-20). Written informed consent was obtained from all participants. We used modified respondent-driven sampling (RDS) technique [19] to recruit IDPWID in Odessa, Ukraine, in June through September 2020. We started recruitment with four index cases, who were defined as people who 1) reported IDU in the last 30 days, 2) were internally displaced from Donetsk, Lugansk regions or Crimea because of the military conflict irrespective of their nationality, 3) were living with HIV, but were not receiving ART treatment at the time of study enrolment (Figure 1); and 4) were at least 18 years old. All participants who tested positive for HIV during the study were counselled by trained healthcare workers and linked to care. Unlike in the classic RDS approach, the number of distributed coupons differed depending on the HIV status of a participant: the index cases were given eight coupons to recruit other IDPs with history of IDU (not necessarily recent IDU experience); HIV-positive network members who were not in treatment at the time of the interview were offered up to eight coupons to recruit other IDPWID; HIV-positive IDPWID receiving ART and HIV-negative network members were offered three coupons to recruit other IDPWID. Upon recruitment, participants were briefly interviewed about their socio-demographic characteristics and their migration experience. Recruitment was terminated after the field study period of two months was completed.

**Figure 1.**
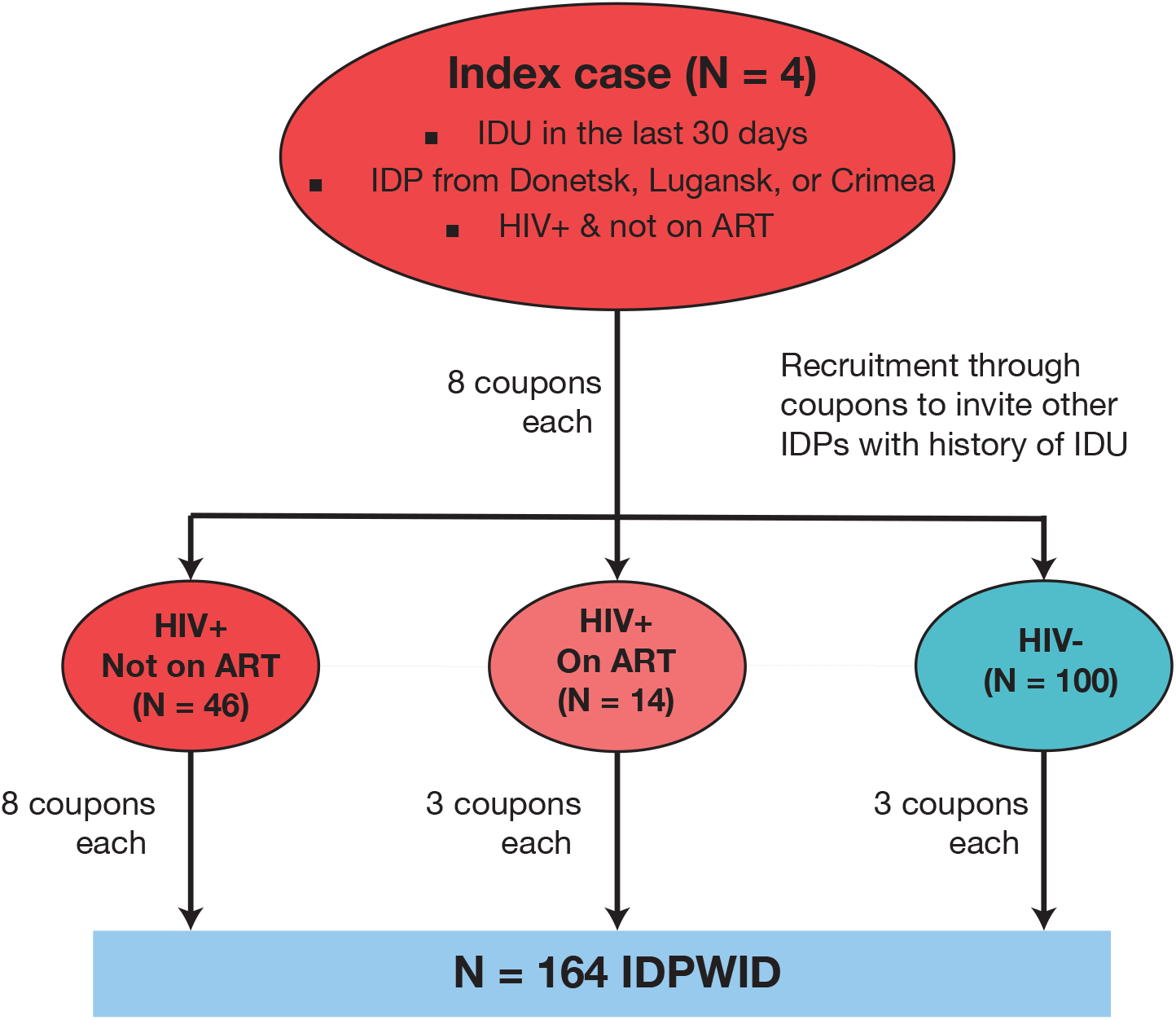
Recruitment flow diagram. IDU – injection drug use, ART – anti-retroviral treatment, IDPs – internally displaced persons, IDPWID – internally displaced people who inject drugs.

### Data Collection

All participants were interviewed about their migration history (e.g. region of origin, date of displacement, date of arrival to Odessa), their socio-demographic and housing information, and then were screened for HIV and HCV by rapid tests (Supplementary Methods). Whole blood samples were collected from all participants, and serum was isolated at the Odessa Regional Virology Laboratory and stored at ─80°C.

### Statistical analysis

We used chi-square tests to investigate whether participants’ recent injection drug use, HIV and HCV status were significantly associated with their housing status (Supplementary Table 3). We defined current IDU status as IDU in the last 30 days (yes/no). The result of rapid HIV and HCV test performed in this study was used to define HIV and HCV status. We dichotomized the housing status into stable and unstable. We defined stable housing as living in own house or flat, living with relatives or friends, and living in rented accommodation; unstable housing was defined as living in IDPs shelters, frequently changing accommodation, being homelessness and other housing types for example hostels and rehabilitation centres.

### HCV genome sequencing

Although MinION based HCV sequencing methods have been described previously [20], we aimed to develop an alternative protocol that meets the clinical need for generation of near real-time sequencing data in field settings, without compromising sample integrity or losing sensitivity. Methods were adapted from the ARTIC Network nCoV-2019 Sequencing Protocol V3 LoCost (https://www.protocols.io/view/ncov-2019-sequencing-protocol-v3-locost-bh42j8ye). Briefly, we employed a 400 bp tiling amplicon scheme with genotype- and subtype-specific primers designed to generate partial or NFLG HCV sequences as per the Primal Scheme protocol using reference strains for HCV genotypes 1a, 1b, and 3a (see Supplementary Methods and Supplementary Tables 1 and 2). The obtained cDNA amplicons were purified, pooled, and used for library preparation with the Ligation Sequencing Kit (SQK-LSK109) (Oxford Nanopore Technologies, Oxford, UK). Final libraries were loaded onto a flow cell (FLO-MIN106) and sequenced with the MinION device. Consensus sequences from this study (Accession Numbers #) have been deposited into GenBank. All sequencing work was carried out in a simulated field environment using a “lab-in-a-suitcase” approach [18], hydroponic grow tents were implemented to physically separate pre-PCR, post-PCR, and master mix work to prevent sample contamination (Meredith et al., unpublished). Further details including bioinformatic workflow, beyond the description in this section, are presented in the Supplementary Materials.

### Phylogenetics

HCV sequences were only included in further phylogenetic analysis if the obtained consensus sequence covered >=50% of the reference genome (sensitivity analyses were performed with other coverage levels and are presented in the Supplementary materials). For each identified HCV subtype, all resulting consensus genome HCV sequences were aligned using the Muscle algorithm in AliView [21]. RaxML [22] was then used to reconstruct maximum likelihood (ML) phylogenetic trees for each subtype under a general time-reversible nucleotide substitution model with gamma-distributed rate-variation among sites (GTR+Γ). We used ClusterPicker [23] to identify possible transmission clusters defined as clades with >90% bootstrap support and within-clades genetic distance <3%. Other previously published cluster-defining criteria were considered [14], but did not affect our findings (Supplementary Materials).

We used BEAST 1.10.4 [24] to perform molecular clock analyses and reconstruct population growth history for each of the identified HCV subtypes. We used the Bayesian Skyline population growth model (10 intervals), the GTR+Γ nucleotide substitution model, and the uncorrelated lognormal relaxed clock model. Since our datasets did not have enough molecular clock signal (estimated by TempEst [25], Supplementary Table 4), we used previously published estimates for evolutionary rates of HCV subtypes to inform our analyses. Specifically, we used normally distributed priors with mean = 1.48E-3 and standard deviation = 2.3E-4, mean = 1.18E-3 and standard deviation = 2.1E-4, mean = 1.289E-3 and standard deviation = 1.47E-4, for subtypes 1a, 1b, and 3a, respectively [26, 27].

## Results

### Study population

A total of 164 IDPWID were recruited in Odessa, Ukraine, between June through September 2020 (Table 1). All participants had complete data, apart from one participant for the variable “previous HCV test results” which is indicated in Table 1. For each variable, cases with missing data were eliminated. Most participants identified as male (81.1%), completed secondary or higher education (73.2%), were single (46.9%), reported recent IDU (92.7%), and arrived in Odessa between 2014-2015 (53.7%). Six participants reported ever engaging in sex work and two participants reported men who have sex with men (MSM) contacts. The median participants’ age was 37 years old (range 20 – 63) at the time of enrolment. The median length of IDU experience was 10 years (range 2 – 40).

**Table 1.**
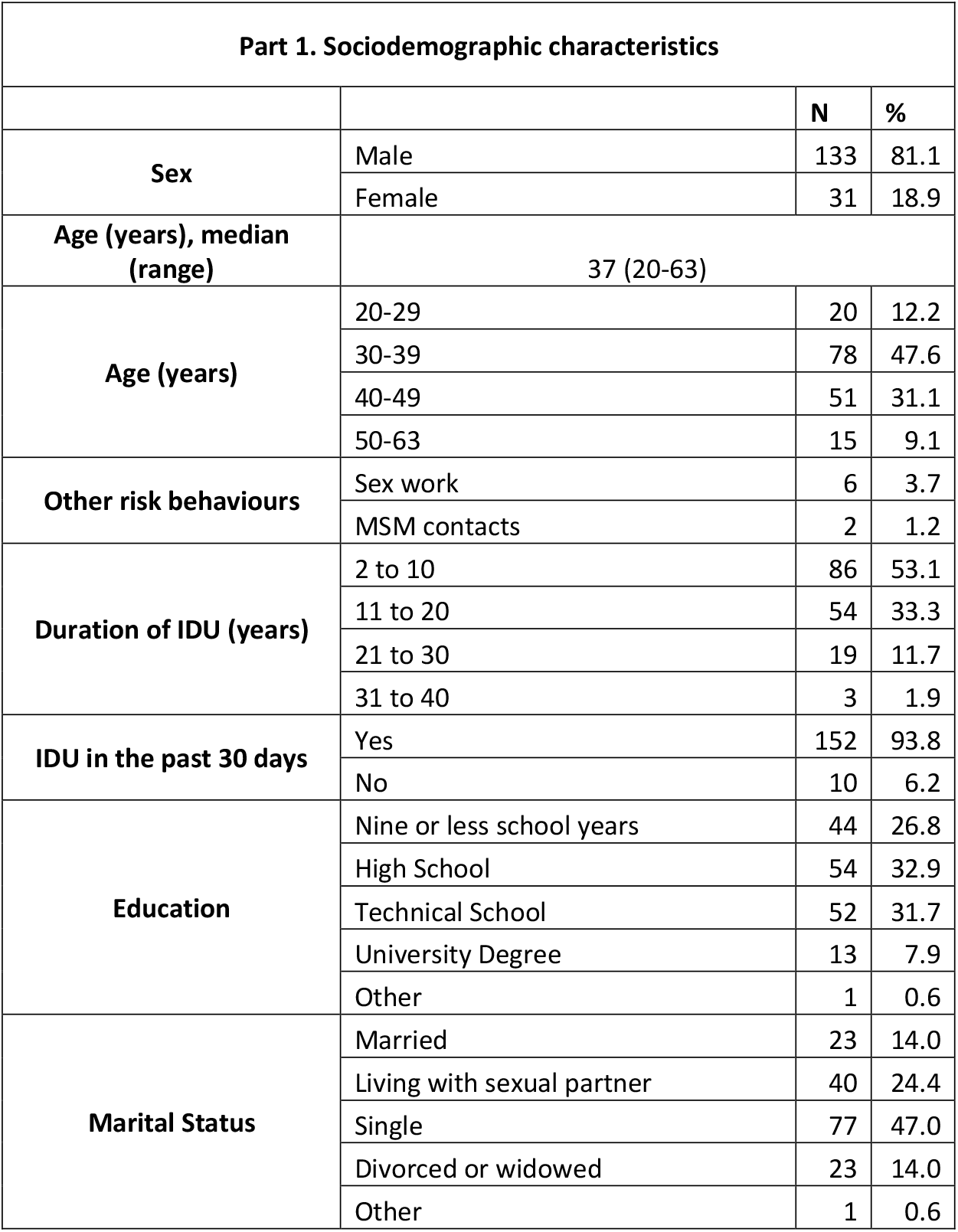
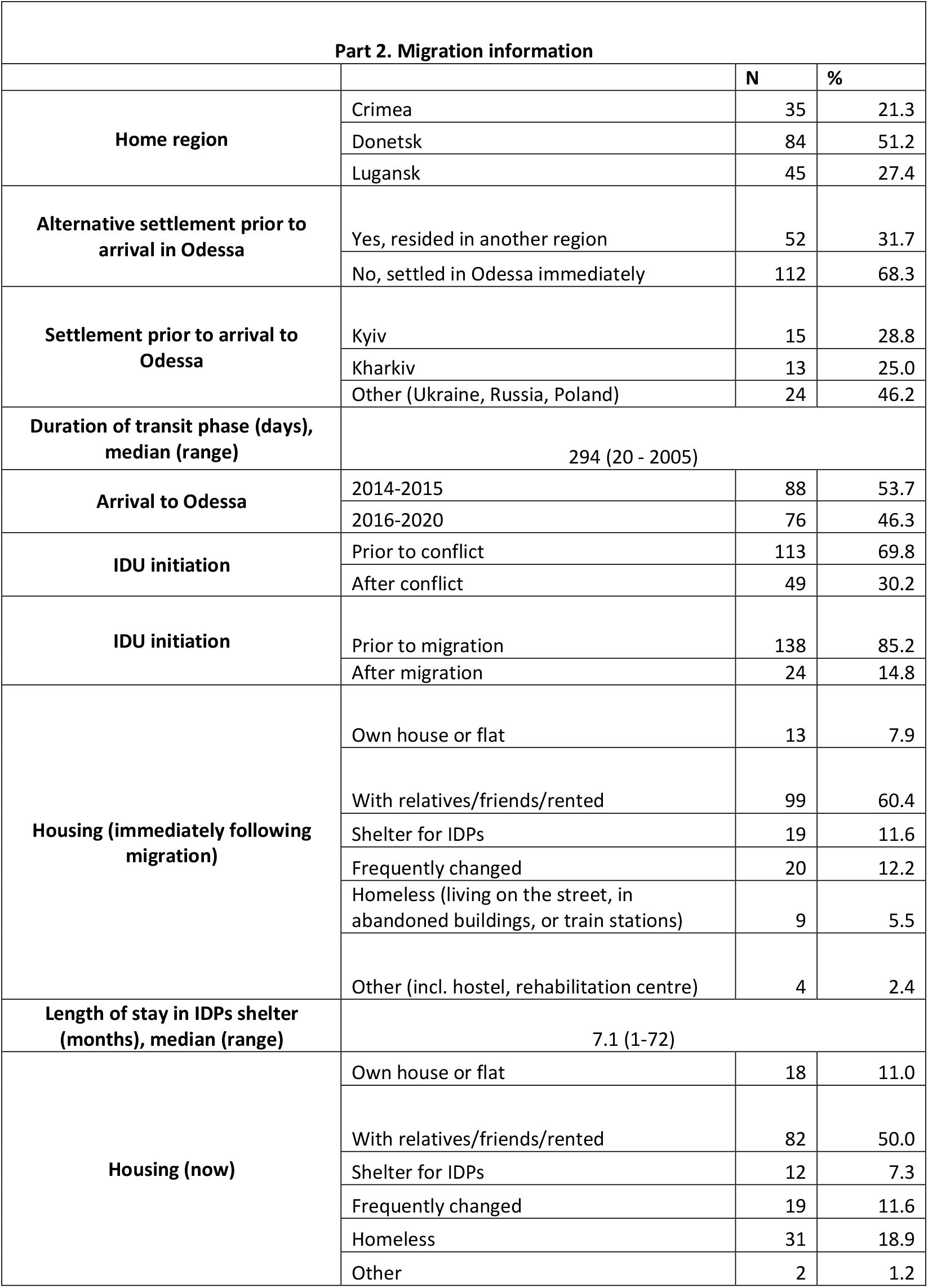
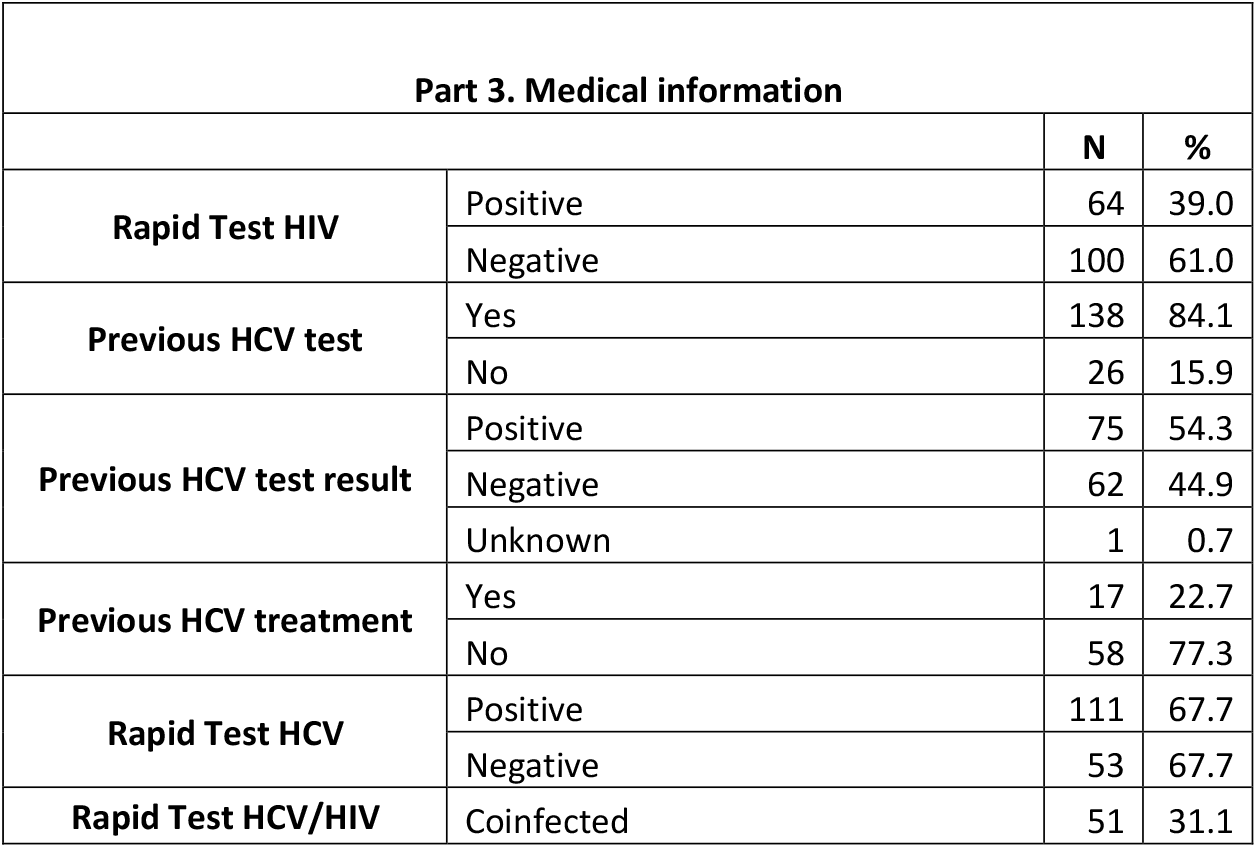
Characteristics of IDPWID recruited in Odessa in 2020

| Part 1. Sociodemographic characteristics |  |  |  |
| --- | --- | --- | --- |
|  |  | N | % |
| Sex | Male | 133 | 81.1 |
|  | Female | 31 | 18.9 |
| Age (years), median (range) | 37 (20-63) |  |  |
| Age (years) | 20-29 | 20 | 12.2 |
|  | 30-39 | 78 | 47.6 |
|  | 40-49 | 51 | 31.1 |
|  | 50-63 | 15 | 9.1 |
| Other risk behaviours | Sex work | 6 | 3.7 |
|  | MSM contacts | 2 | 1.2 |
| Duration of IDU (years) | 2 to 10 | 86 | 53.1 |
|  | 11 to 20 | 54 | 33.3 |
|  | 21 to 30 | 19 | 11.7 |
|  | 31 to 40 | 3 | 1.9 |
| IDU in the past 30 days | Yes | 152 | 93.8 |
|  | No | 10 | 6.2 |
| Education | Nine or less school years | 44 | 26.8 |
|  | High School | 54 | 32.9 |
|  | Technical School | 52 | 31.7 |
|  | University Degree | 13 | 7.9 |
|  | Other | 1 | 0.6 |
| Marital Status | Married | 23 | 14.0 |
|  | Living with sexual partner | 40 | 24.4 |
|  | Single | 77 | 47.0 |
|  | Divorced or widowed | 23 | 14.0 |
|  | Other | 1 | 0.6 |

| Part 2. Migration information |  |  |  |
| --- | --- | --- | --- |
|  |  | N | % |
| Home region | Crimea | 35 | 21.3 |
|  | Donetsk | 84 | 51.2 |
|  | Lugansk | 45 | 27.4 |
| Alternative settlement prior to arrival in Odessa | Yes, resided in another region | 52 | 31.7 |
|  | No, settled in Odessa immediately | 112 | 68.3 |
| Settlement prior to arrival to Odessa | Kyiv | 15 | 28.8 |
|  | Kharkiv | 13 | 25.0 |
|  | Other (Ukraine, Russia, Poland) | 24 | 46.2 |
| Duration of transit phase (days), median (range) | 294 (20 - 2005) |  |  |
| Arrival to Odessa | 2014-2015 | 88 | 53.7 |
|  | 2016-2020 | 76 | 46.3 |
| IDU initiation | Prior to conflict | 113 | 69.8 |
|  | After conflict | 49 | 30.2 |
| IDU initiation | Prior to migration | 138 | 85.2 |
|  | After migration | 24 | 14.8 |
| Housing (immediately following migration) | Own house or flat | 13 | 7.9 |
|  | With relatives/friends/rented | 99 | 60.4 |
|  | Shelter for IDPs | 19 | 11.6 |
|  | Frequently changed | 20 | 12.2 |
|  | Homeless (living on the street, in abandoned buildings, or train stations) | 9 | 5.5 |
|  | Other (incl. hostel, rehabilitation centre) | 4 | 2.4 |
| Length of stay in IDPs shelter (months), median (range) | 7.1 (1-72) |  |  |
| Housing (now) | Own house or flat | 18 | 11.0 |
|  | With relatives/friends/rented | 82 | 50.0 |
|  | Shelter for IDPs | 12 | 7.3 |
|  | Frequently changed | 19 | 11.6 |
|  | Homeless | 31 | 18.9 |
|  | Other | 2 | 1.2 |

| **Part 3. Medical information** |  |  |  |
| --- | --- | --- | --- |
|  |  | **N** | **%** |
| **Rapid Test HIV** | Positive | 64 | 39.0 |
|  | Negative | 100 | 61.0 |
| **Previous HCV test** | Yes | 138 | 84.1 |
|  | No | 26 | 15.9 |
| **Previous HCV test result** | Positive | 75 | 54.3 |
|  | Negative | 62 | 44.9 |
|  | Unknown | 1 | 0.7 |
| **Previous HCV treatment** | Yes | 17 | 22.7 |
|  | No | 58 | 77.3 |
| **Rapid Test HCV** | Positive | 111 | 67.7 |
|  | Negative | 53 | 67.7 |
| **Rapid Test HCV/HIV** | Coinfected | 51 | 31.1 |

### Migration and IDU

Most IDPs were originally residing in the Donetsk region (51.2%) and migrated within a year of the beginning of the military conflict in April 2014 (69.5%). Most immediately settled in Odessa following migration (68.3%); of the rest, common regions for intermediate settlement prior to arrival in Odessa included Kyiv (28.8%) and Kharkiv (25%) amongst other locations across Ukraine, Russia, and Poland (Figure 2). For those who settled in Odessa immediately, the reported transit phase between leaving the region of origin and arriving to Odessa was two days or less, whilst for those who had settled elsewhere prior to arrival in Odessa, the median transit phase was 294 days (range 20 days - 5.5 years). In this study, 30.2% of IDPWID did not inject drugs prior to the conflict, of whom 14.8% reported starting IDU upon arrival to Odessa.

**Figure 2.**
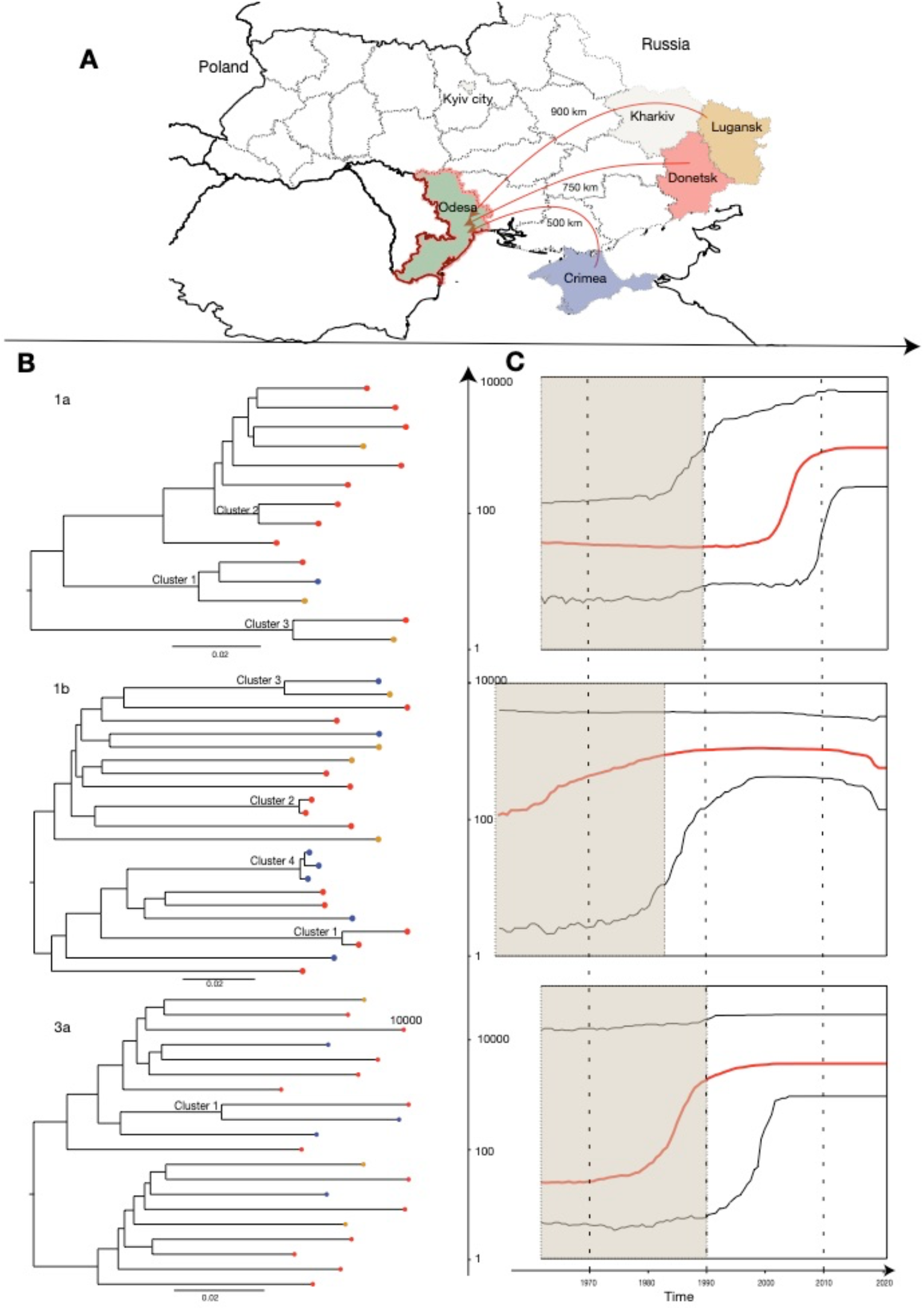
A – Map of Ukraine illustrating migration journeys. Beige colour indicates regions that were often a place of settlement prior to arrival to Odessa. B – Maximum Likelihood phylogenetic trees reconstructed for each HCV genotype and phylogenetic clusters marked on the trees. Tree tips colours correspond to the home regions of IDPWID. Scale bars indicate substitutions/site/year. C – Bayesian Skyline plots reflecting effective population size over time estimated for each of the identified HCV subtypes (red horizontal line – median, black horizontal lines – 95% HPD). Black dotted lines represent the median and lower 95% HPD bound of the time to most recent common ancestor for each subtype. Y-axis indicates log effective population size.

### HIV and HCV prevalence

Previous HIV and HCV testing was reported by 91.5% and 84.1% of participants, respectively. Rapid testing identified seroprevalence of HIV, HCV, and HIV/HCV coinfection of 39%, 67.7%, and 31.1%, respectively. Of those who previously reported a positive HCV test (54.3%), 22.7% received treatment for HCV infection in the past. No participants reported COVID-19 symptoms (high temperature, a new cough, loss of taste or smell in the last 21 days) at the time of interview.

### Migration and housing

The majority of IDPWID in our sample settled with relatives, friends, or in rented accommodation, both immediately following migration and at the time of the study in 2020 (50% and 60.4%, respectively). Of those residing in an IDPs shelter immediately following migration (11.6%), the median length of stay was seven months (range one month - six years). Over three times more participants reported homelessness (living on the street, in abandoned buildings, or train stations) in Odessa at the time of the study compared to immediately following migration (5.5% and 18.9%, respectively). There was no difference between those reporting stable (own house, staying with relatives/friends, or in a rented house) and unstable (staying in the IDPs shelter, changing accommodation frequently, or being homeless) housing at the time of study enrolment in terms of their HIV and HCV statuses (Supplementary Table 3). However, unstable housing amongst IDPWID at the time of the study enrolment was associated with IDU use in the last 30 days (p=0.048).

### HCV sequencing data

Of the 164 people recruited, 90 samples resulted in PCR amplification producing 57 samples with partial (>50%) or NFLG sequences, and 33 samples with sequence coverage <50%. Samples with <50% coverage may be due to low sample quality, low viral loads, as well as low specificity of primer design and availability of reference genomes. These samples were not included for further analysis and future amplification methods will need to be adapted to improve amplification success rates. Of the samples that failed to amplify (74 or 45.1%), 35 were HCV antibody-negative by rapid test, and 39 were HCV antibody-positive; the latter may indicate successful treatment or spontaneously cleared infections. Of the 57 partial or NFLG HCV sequences generated (34.8% of the total 164 samples), genotypes 1a (N = 14), 1b (N = 23), and 3a (N = 20) were identified. Of these, 31 (54.4%) had previously tested positive for HCV but had not received treatment, and of whom five had received a positive anti-HCV test before moving to Odessa, which may indicate chronic infections or re-infection following spontaneous clearance. A total of four participants (7.0%) previously tested positive for HCV and received treatment in the past. Further treatment information would be necessary, which was unavailable in this study, to interpret whether the infection was not cleared, or whether this is a re-infection. At least 22 of these 57 participants were newly diagnosed with HCV in this study: 11 of whom tested negative before and 11 were not tested before (although this does not take into consideration the four participants who received prior treatment as we were unable to distinguish re-infections).

### HCV phylogenetics and phylodynamics

Phylogenetic trees were reconstructed separately for subtypes 1a, 1b, and 3a and included 14, 23, and 20 sequences, respectively (Figure 2). We identified three, four, and one potential transmission clusters (six clusters of two sequences, and two clusters of three sequences) on the subtype 1a, 1b, and 3a trees, respectively. Phylodynamic analysis showed that the time of most recent common ancestor (TMRCA) was in 1958 (95% Highest Posterior Density (HPD) 1915 - 1990), 1949 (1897 - 1983), and 1955 (1907 - 1992) for subtypes 1a, 1b, and 3a, respectively. The reconstructed Bayesian Skyline plots showed that the exponential growth of different HCV subtypes likely started in different decades (Figure 2), with subtype 1b growing before the 1990s, subtype 3a growing before the 2000s (coinciding with the HIV epidemic growth), and subtype 1a before the 2010s. The limited number of sequences in our analyses result in wide confidence intervals and prevent further interpretations of the past epidemic growth history. The TMRCAs for all subtype 1a clusters, subtype 1b cluster 6, and the 3a cluster dated pre-conflict and were estimated to be between 1987 and 2013, unlikely capturing a recent transmission event (Table 2 and Figure 3). Three of the subtype 1b clusters were more recent: TMRCAs for cluster 4, 5, and 7 were estimated to be in November 2011 (May 2005 – April 2016), May 2018 (August 2016 - October 2019), and December 2017 (October 2015 - May 2019), respectively.

**Table 2.** Cluster composition and time to the most recent common ancestor (TMRCA) for HCV phylogenetic clusters

| **Subtype** | **Cluster** | **Number of sequences** | **TMRCA** | **TMRCA 95% HPD** | **Time of IDPWID arrival to Odessa** | **Home region** | **Rapid HIV test result** |
| --- | --- | --- | --- | --- | --- | --- | --- |
| 1a | 1 | 3 | 1998 | 1982 - 2009 | 2014, 2014, 2016 | Lugansk, Donetsk, Crimea | Pos, Pos, Pos |
|  | 2 | 2 | 2005 | 1993 - 2013 | 2015, 2017 | Donetsk, Donetsk | Pos, Neg |
|  | 3 | 2 | 2000 | 1984 - 2011 | 2016, 2015 | Donetsk, Lugansk | Pos, Pos |
| 1b | 4 | 2 | Nov 2011 | May 2005 - April 2016 | May 2015, Jun 2015 | Lugansk, Donetsk | Pos, Pos |
|  | 5 | 2 | May 2018 | Aug 2016 - Oct 2019 | Aug 2014, Mar 2018 | Donetsk, Donetsk | Pos, Pos |
|  | 6 | 2 | 1999 | 1982 - 2010 | 2018, 2017 | Lugansk, Crimea | Neg, Neg |
|  | 7 | 3 | Dec 2017 | Oct 2015 - May 2019 | Jun 2015, Sep 2017, Oct 2015 | Crimea, Crimea, Crimea | Neg, Neg, Neg |
| 3a | 8 | 2 | 1987 | 1961 - 2004 | 2018, 2018 | Crimea, Donetsk | Neg, Pos |

**Figure 3.**
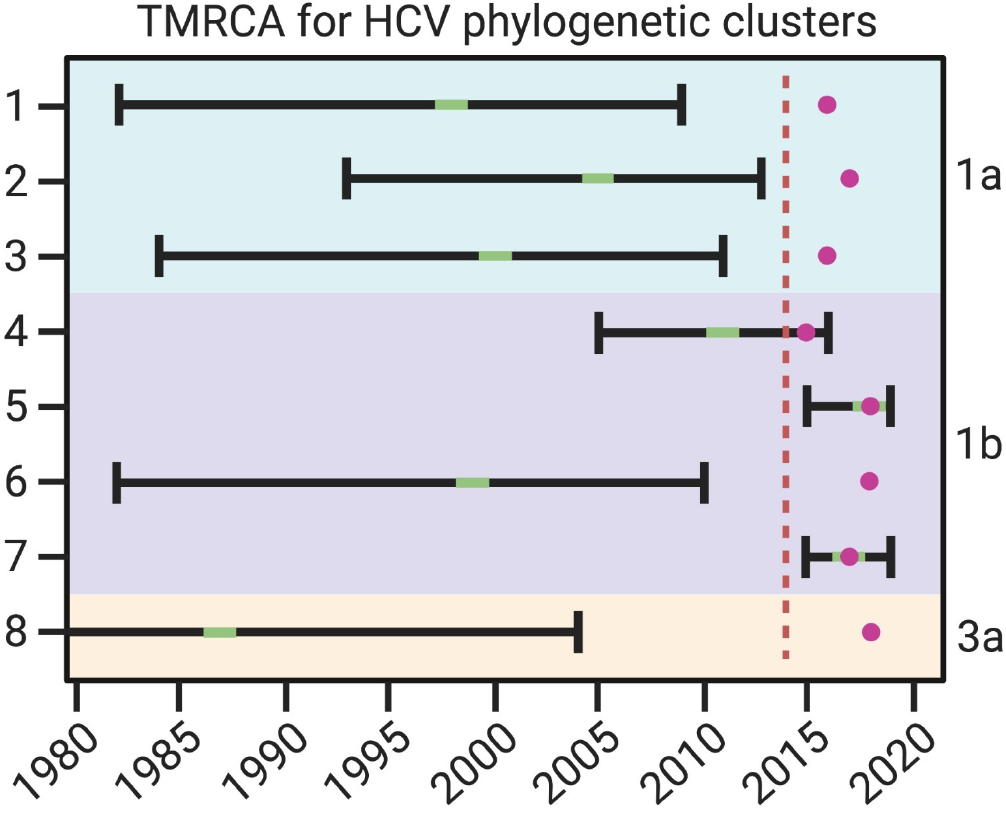
TMRCA uncertainties for HCV phylogenetic clusters in relation to conflict and migration timings. Green indicates TMRCA and black bars are 95% HPD. Purple circle indicates time of latest IDPWID arrival to Odessa within that cluster. Red dashed line indicates the start of the conflict.

## Discussion

Here, we use Nanopore technology to obtain the first HCV genomes from Ukraine and show that Nanopore technology can be used to sequence HCV in a field simulated environment and that molecular clock analysis allows us to estimate timing of viral transmission events relative to the timing of forced migration. Specifically, we show evidence that HCV transmission events are likely to occur soon after arrival to the host region. Of the 57 participants from whom partial or NFLG HCV sequences were available, 29% were found in a total of eight potential infection clusters. Two of these clusters originated after members moved to Odessa: in both cases, the HCV transmission event happened within at most one and a half years upon arrival of the last individual to the city, suggesting that prevention efforts in long-term settlement environments will be beneficial soon after migration. In both of these recent clusters, although transmissions happened after migration, all cluster members originated from the same home regions (Donetsk and Crimea for clusters 4 and 7, respectively), which supports previous observations of the importance of social networks from the same home community post-migration amongst forcibly displaced people [28]. The clusters we identified are an underestimation of the true number of HCV transmissions in this group and are only indicative of transmission within the IDPWID community. Absence of other published HCV genomes from Ukraine made it impossible to compare clustering within and between migrant, home, and host communities.

Previous reports of genotype distribution in the general population in Ukraine in the early 2010s showed low prevalence of HCV infection with subtype 1a (1.6%), and a high prevalence of 1b (42.1%), followed by 3 (28.8%) [9]. In this study, we report much higher prevalence of HCV infection with the 1a (25%) subtype, and comparable levels of infection with 1b (40%) and 3a (35%) within IDPWID in Odessa. The HCV subtype 1b dominates epidemics in the neighboring Russian Federation (1a – 2.1%; 1b – 52.8%; 3 – 36.3%) and several post-Soviet countries, such as Uzbekistan and Azerbaijan, whilst 1a is more commonly found in western Europe [9, 29]. Higher 1a prevalence amongst HIV co-infected patients compared to the general population has been linked to IDU mediated transmission elsewhere [30], and thus is expected in our study population. The increase in 1a prevalence in our study, reflected in the most recent growth of this subtype compared to subtypes 1b and 3a as estimated by our reconstructed Bayesian Skyline plots, follows the increase in HIV transmission in PWID [31].

Our results suggest that HCV seroprevalence in IDPWID currently residing in Odessa (67.7%) is now higher than the prevalence reported in 2015 in both autochthonous PWID (47.5%) and in PWID residing in Donetsk (55.2%) [10]. This may reflect our recruitment strategy, given the enhanced coupon distribution for HIV-positive participants who were not in ART treatment at the time of enrolment. Also, 22% of those receiving a positive anti-HCV test result had also received treatment in our study, compared to only 9.4% in a 2013 study, which may be due to a recent increase in targeted HCV treatment efforts for the PWID community [32, 33]. We generated four HCV genomes from IDPWID who previously received treatment, of which two were found in potential transmission clusters, likely indicating a re-infection or an incomplete treatment course. Continued engagement in treatment is crucial for transmission and disease progression prevention, but forcibly displaced people might face additional barriers in accessing these services [3, 4].

Due to a limited sample size, we were unable to link epidemiological data with our clusters, however we made important observations which can help inform targeted public health strategies. We report an increase in homelessness, an independent risk factor for HIV and HCV acquisition for PWID [6], at the time of the study (18.9%), compared to the period immediately following migration (5.5%). Most of those who lost their homes were residents of IDPs shelters or stayed with relatives/friends or rented their own accommodation upon arrival. Unstable housing at the time of the study was correlated with more IDPWIDs engaging in IDU in the last 30 days. We acknowledge the complexity of the categorisation of housing types in the literature, which may classify living with relatives/friends as unstable housing. In this study, we included the option of “frequently changing accommodation” which we believe may help encompass some of the hardships and risk factors faced when living with relatives/friends and which are characteristic of others in the unstable category. As such, the option of living with relatives/friends/rented was included in the stable category. Overall, IDPs are likely to reside in high-risk environments with limited prevention and treatment resources, thus long-term housing solutions for IDPs are likely to help minimise infection risks [4].

Many IDPWID in our sample initiated IDU after the beginning of the conflict (30.2%) or even after they migrated to Odessa (14.8%). Substance use amongst forcibly displaced people as a coping mechanism following conflict, violence, and migration stress is well documented in current literature [34]. In our study, two individuals who reported starting IDU after migration were found in HCV transmission clusters with those reporting 15 to 20 years IDU experience. Although IDPWID with prior experience of IDU may be more likely to seek and engage with harm reduction programs, efforts to inform and engage new injectors immediately following migration should be prioritised to reduce HCV transmission amongst this population.

Forced migration is a highly heterogenous and complex phenomenon, and is dependent on multiple factors including historical, economic, social, and political landscapes. The migration journey may differ across time: exposure to risk factors and hardships may continuously evolve. Regions with large migrant populations, limited laboratory capacities, and high prevalence of infectious diseases can especially benefit from the proposed integrative phylogenetic and epidemiological approach. Increased monitoring and understanding of the dynamics of infectious disease transmission networks, timing of transmission events, and risk factors unique to the experiences faced by forcibly displaced people is a crucial step in the development of effective health interventions.

## Supporting information

STROBE Checklist Cross Sectional

Coi Disclosure

## Data Availability

All data produced in the present study are available upon reasonable request to the authors

## Acknowledgements

We acknowledge the assistance of public health workers from the non-governmental organisations “Alliance of Public Health” and “Way Home” in collection of epidemiological and behavioural data and biological samples, and their ongoing efforts of providing preventative measures and infection management to at risk populations in Ukraine. We also express our sincere gratitude towards study participants. We thank Rowena Bull and Chaturaka Rodrigo for their advice with MinION based hepatitis C sequencing protocols. This work was funded by the John Fell Fund and the Wellcome Institutional Strategic Support Fund at the University of Oxford. TIV is supported by the Branco Weiss Fellowship. JOW is supported by an NIH-NIAID R01 (AI135992). Bioinformatics analyses reported herein were in part supported by an NIH NIGMS Institutional Development Award (IDeA), grant no. P20GM103395 (Alaska INBRE: Pathogenomics server – to E.B.). A.Y. was supported by a University of Cambridge BBSRC DTP studentship at the time of the study. IG is a Wellcome Senior Fellow. GK, LM, and IG are supported by grants from the Wellcome Trust (Refs: 207498/Z/17/Z and 206298/B/17/Z).

## Supplementary Methods

### Study Design and Data Collection

Participants were interviewed by social workers from the non-governmental organisation “Way Home”. At the time of interview, all participants were screened for HIV and HCV using Wondfo® One Step HIV1/2 Whole Blood/Serum/Plasma and Wondfo® One Step HCV Whole Blood/Serum/Plasma.

### RNA Extraction and cDNA Synthesis

Viral RNA was extracted from 140 μL serum using QIAamp Viral RNA Minikit (Qiagen), following the manufacturer’s instructions. Extracted RNA was eluted in a 60 μL volume, aliquoted and stored until use at ─80°C. Extracted RNA was reverse transcribed using SuperScript IV Reverse Transcriptase (SSIV RT, ThermoFisher Scientific). Each reaction included 1 μL random hexamers (ThermoFisher Scientific), 1 μL 10 mM dNTP (ThermoFisher Scientific), 3 μL SSIV buffer, 0.5 μL 0.1M DTT, 0.5 μL RNaseOUT (ThermoFisher Scientific), 0.5 μL SSIV RT and 6.5 μL RNA extract. The mix was incubated at 23°C for 10 min, 50°C for 50 min, 75°C for 10 min, and then held at 4°C.

### Primer Design

To amplify and sequence low abundance viruses directly from the clinical samples we employed a tiling amplicon scheme with the genotype- and subtype-specific primers. Primers were designed to generate near full length HCV genome sequences as per the Primal Scheme (https://primalscheme.com) protocol based on multiplex tiling PCR amplification of 400bp amplicons overlapping neighbouring amplicons by 75 nt [1]. A collection of reference strains for HCV genotypes 1a, 1b, and 3a were identified based on closest available geographical and temporal sequences from GenBank (Supplementary Table 1). Primers for each genotype were pooled into 100 μM pool A and B stock mixes based on alternate regions, which do not overlap within the same pool (Supplementary Table 2).

### PCR Amplification

HCV PCR amplification and sequencing methods were adapted from the ARTIC Network nCoV-2019 Sequencing Protocol V3 LoCost (https://www.protocols.io/view/ncov-2019-sequencing-protocol-v3-locost-bh42j8ye). Briefly, PCR amplifications were performed using Q5 Hot Start High-Fidelity DNA Polymerase (NEB). Two separate reactions for primer pool A and B were prepared in a final volume of 12.5 μL, containing 2.5 μL 5X Q5 Reaction Buffer, 0.25 μL Q5 Hot Start DNA Polymerase, 10 μM primer pool (A or B), 0.5 μL 10 mM dNTPs, 3.75 μL nuclease-free water and 3.75 μl cDNA template. Thermocycling was performed using a mini16 thermal cycler (minipcr) with the following program: 98°C for 30 secs, then 35 cycles of 98°C for 15 secs, 62.5°C for 5 min, and final hold at 4°C. Pool A and B reaction products for a given sample were pooled together in equal volumes and quantified QuantiFluor® ONE dsDNA Dye (Promega) using the Quantus Fluorometer (Promega).

### Oxford Nanopore Library Preparation and Sequencing

Samples were purified with SPRI beads (x1 ratio) and eluted in 25 μL of nuclease-free water. Amplicon ends were prepared for ligation with the NEBNext Ultra II End Repair/dA-Tailing Module (NEB). Unique barcodes from the Oxford Nanopore Native Barcoding Expansion kits EXP-NBD104 (1-12) and EXP-NBD114 (13-24) were ligated to each sample using the NEBNext Ultra II Ligation Module (NEB). Following barcoding, reactions were pooled together in equal volumes and purified with SPRI beads (x0.4 ratio). Finally, Oxford Nanopore sequencing adapters were ligated with the NEBNext Quick Ligation Module (NEB) and the library was purified with SPRI beads again (x1 ratio) and quantified using the Quantus Fluorometer (Promega). Final libraries were loaded onto a flow cell (FLO-MIN106) and sequenced with the MinION.

### Bioinformatic Workflow

All runs were basecalled using guppy basecaller 4.4.2 with the high accuracy model. Demultiplexing was performed as a separate step with guppy barcoder using additional arguments to detect mid strand barcodes. During the demultiplexing step barcodes and an additional 25nt were trimmed from the reads to remove primers. Reads were aligned to 54 HCV reference genomes with Minimap2 [2] and the reference with the highest number of primary mappings as determined by samtools [3] was used as a draft genome for a reference guided assembly. Racon 1.4.21 [4] was used to polish the reference genome with the no trim argument enabled and window size set to 300nt. The draft generated by racon was then polished with medaka consensus (https://github.com/nanoporetech/medaka) and any nucleotides with less than 20x coverage were masked. To test for the presence of multiple genomes within the sample, reads that mapped to the previously generated assembly were removed and the remaining reads were again aligned to the 54 reference genomes. Barcodes with over 200k reads mapped to a second genome were used to generate additional assemblies.

### Sensitivity analysis

Previously published cluster-defining criteria were considered [5, 6]. For subtype 1a analysis, the number of identified clusters was the same if the genetic distance threshold ranged between 3-6%. If the bootstrap support was relaxed to 80 instead of 90 and genetic distance threshold was considered at 4.5%, then 1 additional cluster of 4 sequences was identified. For subtypes 1b and 3a analyses, the same clusters were identified with genetic distance threshold varying 3-6% and bootstrap support of 80.

## Supplementary Tables

**Supplementary Table 1.** Accession numbers for reference genomes used for HCV primer design

| Genotype | Accession number | Genotype | Accession number |
| --- | --- | --- | --- |
| 1a | EU260395 | 1b | GU451218 |
| 1a | D10749 | 3a | AB792683 |
| 1a | KC844049 | 3a | KC844041 |
| 1a | EU482876 | 3a | MG428679 |
| 1a | EU256050 | 3a | GQ275355 |
| 1a | EU239715 | 3b | KC844044 |
| 1a | EU256033 | 3a | JQ717258 |
| 1a | EU256027 | 3a | JQ717256 |
| 1a | EU239713 | 3a | JQ717259 |
| 1a | EU256040 | 3a | JN714194 |
| 1a | EU255932 | 3a | KF035123 |
| 1a | EU155310 | 3a | KF035127 |
| 1a | EU155287 | 3a | KF035126 |
| 1a | EU255951 | 3a | KF035124 |
| 1a | EU256048 | 3a | JQ717260 |
| 1a | EU256097 | 3a | JQ717254 |
| 1a | EU155275 | 3a | JQ717257 |
| 1a | EU155239 | 3a | JQ717255 |
| 1a | EU781746 | 3a | KF035125 |
| 1a | EU256105 |  |  |
| 1a | NC_004102 |  |  |
| 1b | AB435162 |  |  |
| 1b | AF139594 |  |  |
| 1b | AB049088 |  |  |
| 1b | DQ071885 |  |  |
| 1b | AJ238799 |  |  |
| 1b | KC844051 |  |  |
| 1b | EU857431 |  |  |
| 1b | AF176573 |  |  |
| 1b | KC844052 |  |  |
| 1b | AB049101 |  |  |
| 1b | AB249644 |  |  |
| 1b | EF032892 |  |  |
| 1b | GU451224 |  |  |
| 1b | GU451223 |  |  |
| 1b | GU451222 |  |  |
| 1b | GU451221 |  |  |
| 1b | GU451220 |  |  |
| 1b | GU451219 |  |  |
| 1b | GU451218 |  |  |
| 3a | AB792683 |  |  |
| 3a | KC844041 |  |  |

**Supplementary Table 2.**
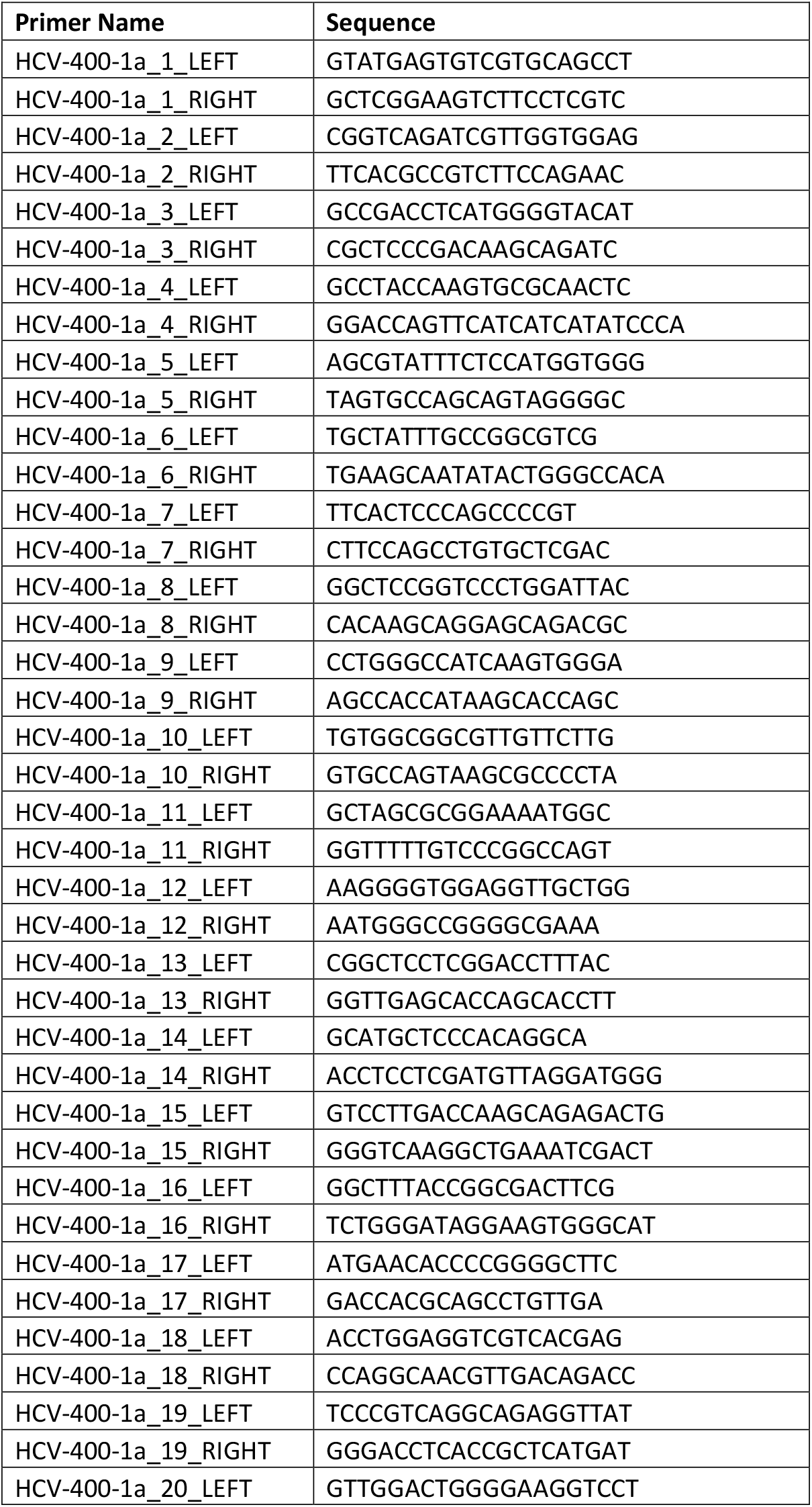
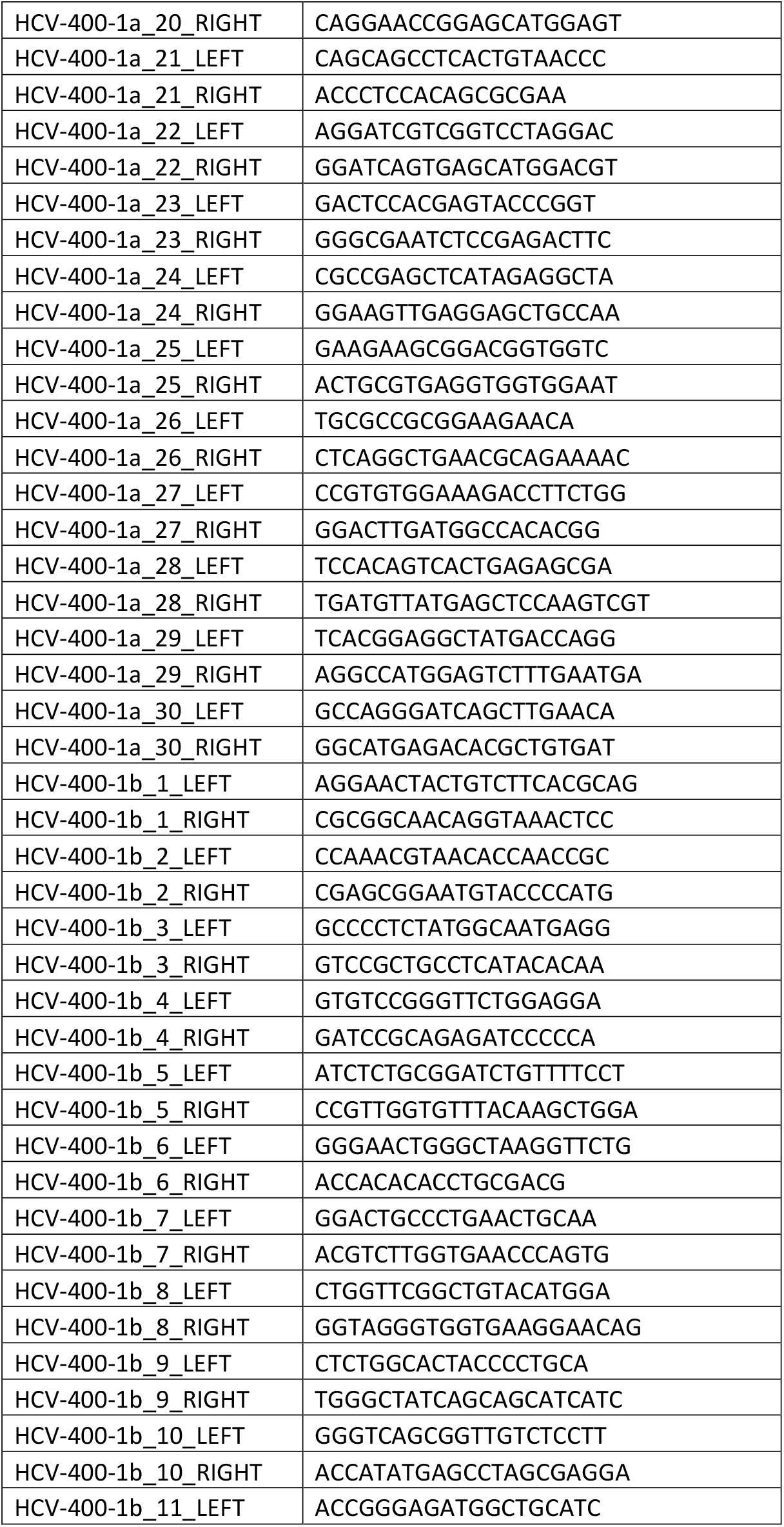
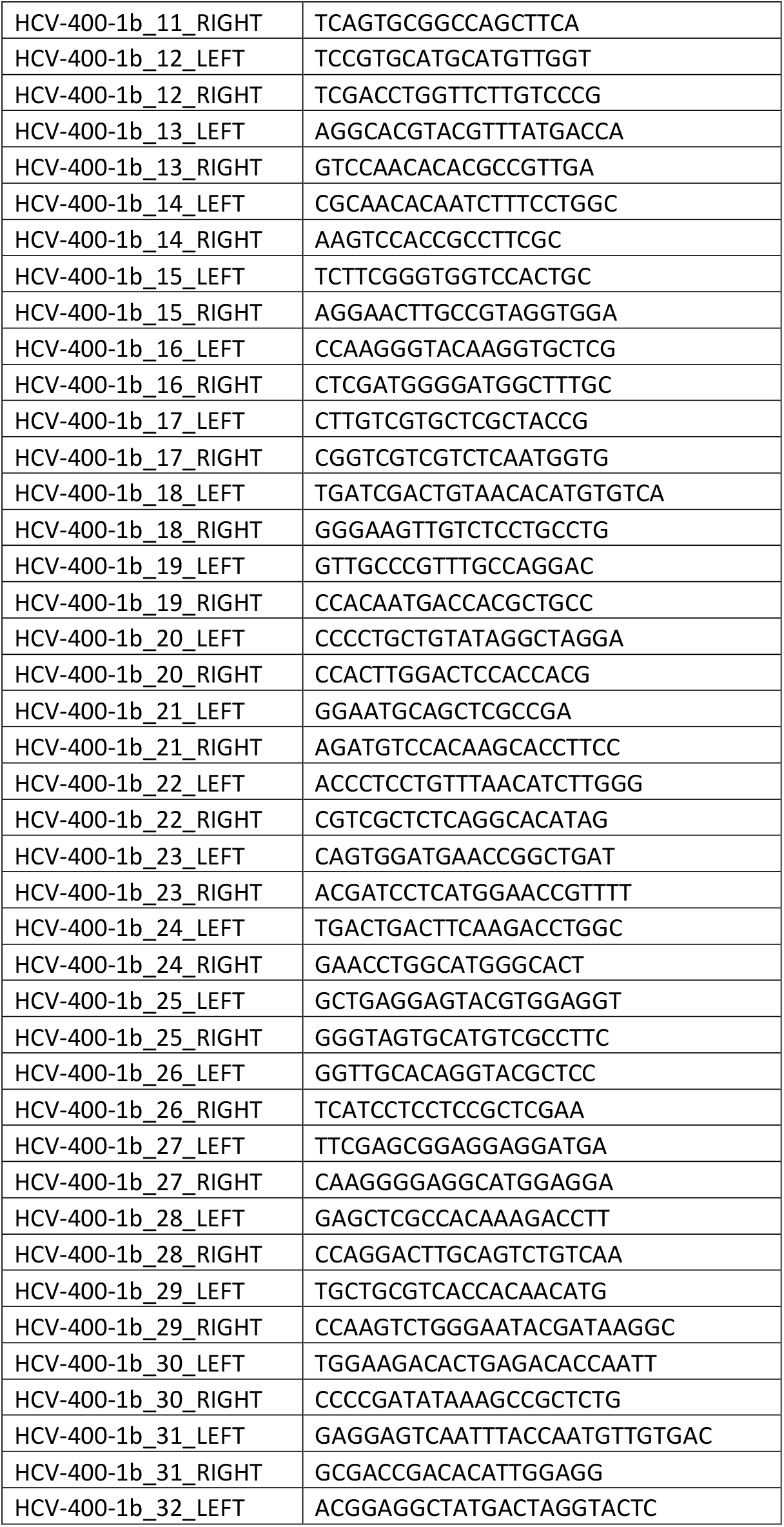
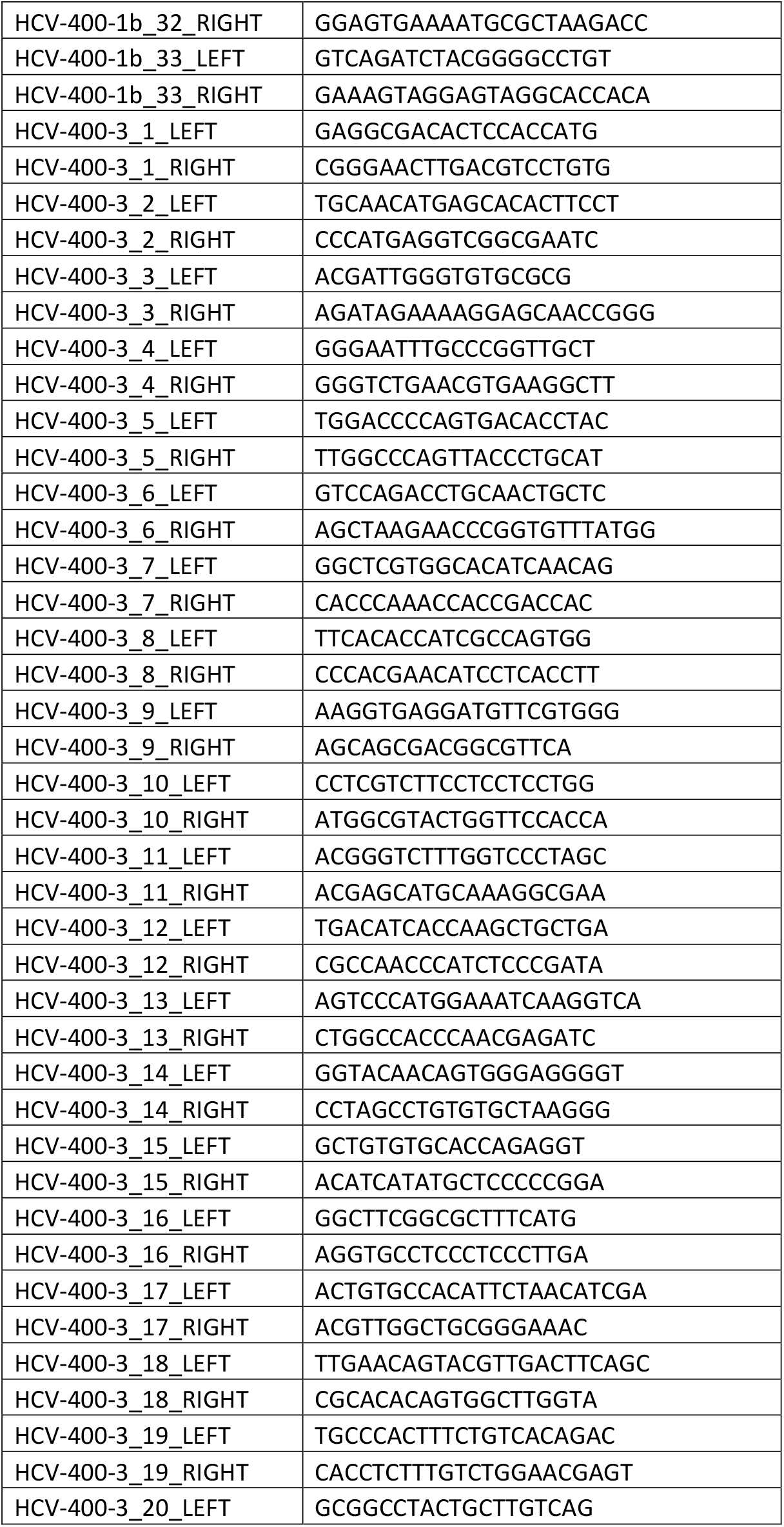
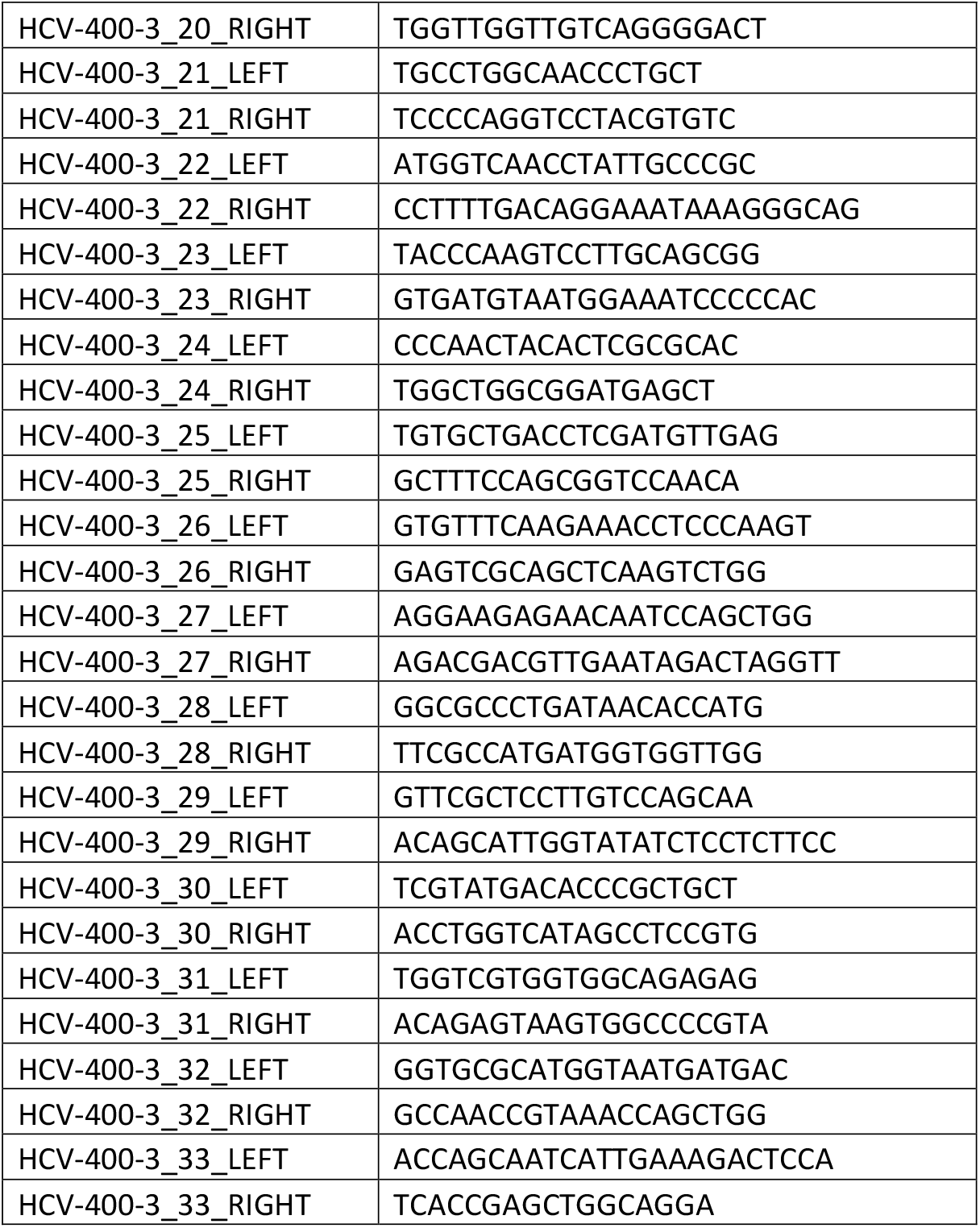
HCV Sequencing Primers

**Supplementary Table 3.** Characteristics of participants based on their housing situation at the time of study enrolment

|  |  | Housing |  |  |
| --- | --- | --- | --- | --- |
|  |  | Stable | Unstable | Chi-square p-value |
| **HIV test result** | Positive | 34 | 30 | 0.1 |
|  | Negative | 66 | 34 |  |
| **HCV test result** | Positive | 71 | 40 | 0.26 |
|  | Negative | 29 | 24 |  |
| **IDU in the past 30 days** | Yes | 89 | 63 | 0.048 |
|  | No | 9 | 1 |  |

**Supplementary Table 4.** Results of the TempEst analysis

| **Dataset** | **Root-to-tip correlation coefficient** | **Estimated evolutionary rate** | **Prior for clock rate** |
| --- | --- | --- | --- |
| 1a | 0.09 | 0.02 | 0.00148 (2.3E-4) |
| 1b | 0.19 | 0.02 | 0.00118 (2.1E-4) |
| 3a | -0.12 | -0.02 | 0.001289 (1.47E-4) |

**Supplementary Table 5.**
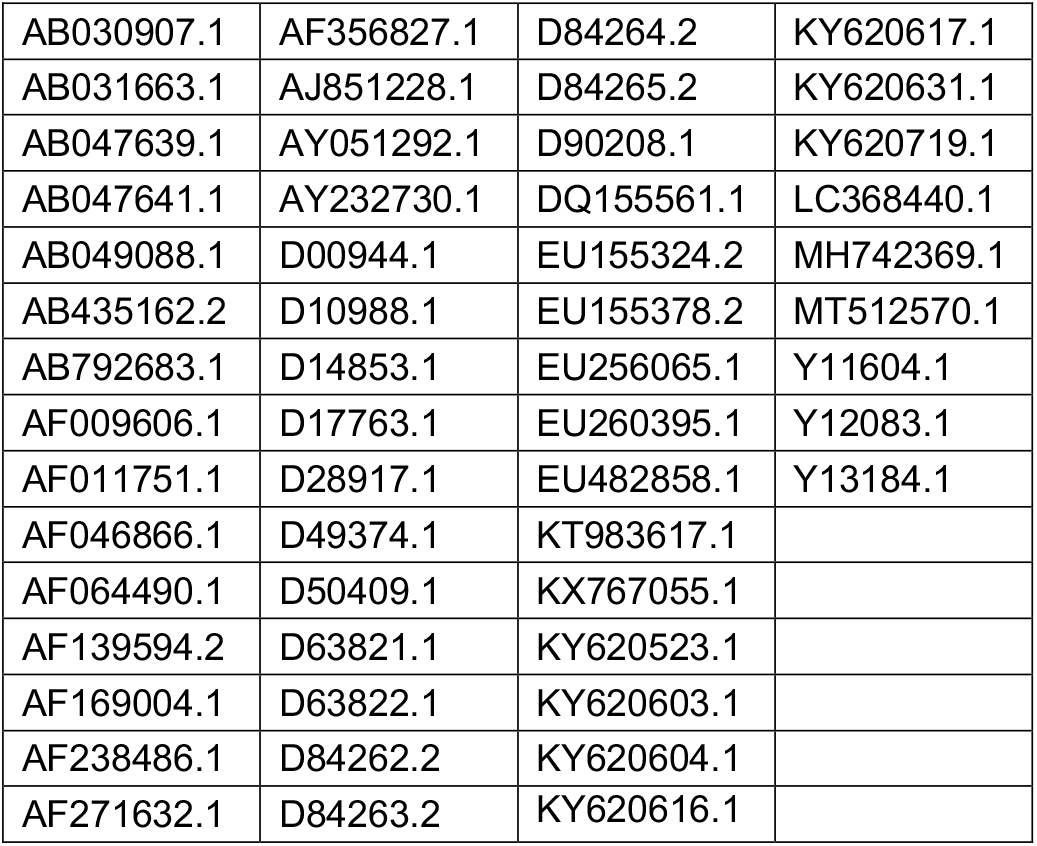
Accession numbers of reference genomes used for HCV genome assembly

|  |  |  |  |
| --- | --- | --- | --- |
| AB030907.1 | AF356827.1 | D84264.2 | KY620617.1 |
| AB031663.1 | AJ851228.1 | D84265.2 | KY620631.1 |
| AB047639.1 | AY051292.1 | D90208.1 | KY620719.1 |
| AB047641.1 | AY232730.1 | DQ155561.1 | LC368440.1 |
| AB049088.1 | D00944.1 | EU155324.2 | MH742369.1 |
| AB435162.2 | D10988.1 | EU155378.2 | MT512570.1 |
| AB792683.1 | D14853.1 | EU256065.1 | Y11604.1 |
| AF009606.1 | D17763.1 | EU260395.1 | Y12083.1 |
| AF011751.1 | D28917.1 | EU482858.1 | Y13184.1 |
| AF046866.1 | D49374.1 | KT983617.1 |  |
| AF064490.1 | D50409.1 | KX767055.1 |  |
| AF139594.2 | D63821.1 | KY620523.1 |  |
| AF169004.1 | D63822.1 | KY620603.1 |  |
| AF238486.1 | D84262.2 | KY620604.1 |  |
| AF271632.1 | D84263.2 | KY620616.1 |  |

